# A causal inference approach for estimating effects of non-pharmaceutical interventions during Covid-19 pandemic

**DOI:** 10.1101/2022.02.28.22271671

**Authors:** Vesna Barros, Itay Manes, Victor Akinwande, Celia Cintas, Osnat Bar-Shira, Michal Ozery-Flato, Yishai Shimoni, Michal Rosen-Zvi

## Abstract

In response to the outbreak of the coronavirus disease 2019 (Covid-19), governments worldwide have introduced multiple restriction policies, known as non-pharmaceutical interventions (NPIs). However, the relative impact of control measures and the long-term causal contribution of each NPI are still a topic of debate. We present a method to rigorously study the effectiveness of interventions on the rate of the time-varying reproduction number R_t_ and on human mobility, considered here as a proxy measure of policy adherence and social distancing. We frame our model using a causal inference approach to quantify the impact of five governmental interventions introduced until June 2020 to control the outbreak in 113 countries: confinement, school closure, mask wearing, cultural closure, and work restrictions. Our results indicate that mobility changes are more accurately predicted when compared to reproduction number. All NPIs, except for mask wearing, significantly affected human mobility trends. From these, schools and cultural closure mandates showed the largest effect on social distancing. We also found that closing schools, issuing face mask usage, and work-from-home mandates also caused a persistent reduction on R_t_ after their initiation, which was not observed with the other social distancing measures. Our results are robust and consistent across different model specifications and can shed more light on the impact of individual NPIs.

## Introduction

The coronavirus disease 2019 (Covid-19) pandemic has caused an enormous impact on the economy and on global public health. As of January 1^st^ 2022, the disease had over 290 million cases and more than 5 million deaths recorded in over 200 countries and territories [1]. In response to the state of emergency declared by the World Health Organization (WHO) in January 2020, governments worldwide have introduced multiple restriction policies, known as non-pharmaceutical interventions (NPI), to mitigate the spread of the severe acute respiratory syndrome coronavirus 2 (SARS-CoV-2). Today, two years from the first outbreak registered in Wuhan China, the relative impact of control measures and the long-term causal contribution of each NPI are still a topic of debate.

In June 2020, Flaxman and colleagues [2] pioneered the challenge of estimating the effectiveness of major interventions on the transmission of SARS-CoV-2. On the basis of mortality data and a Bayesian hierarchical model, they concluded that lockdowns had been effective in most European countries that were studied. Since then, due to the increased widespread adoption of restrictions, several studies have attempted to disentangle the effect of individual NPIs in several countries [3–10] or in the US alone [11–17]. Overall, most previous mathematical modeling suggest that public health interventions were associated with a reduction of Covid-19 incidence. Nonetheless, the conclusions regarding the effect of each specific intervention are not unequivocal. For example, [4, 5, 7, 8, 18] point out that school closures had a significant effect on the transmission of new infections, while a review conducted by [19] suggested that closing schools did not contribute to controlling the pandemic in countries like China, Hong Kong, and Singapore. In fact, existing evidence for the impact of policies is not consistent in the literature, as NPI effectiveness may vary across regions depending on the local context [16].

In this study, we provide a comprehensive analysis of the employment of five NPIs - confinement, school closure, mask wearing, cultural closure, and work restrictions - in 113 countries during the first 5 months of 2020. We observed that during the first-wave period there was a great deal of consistency in the set of restriction measures imposed throughout the world, which was substantially reduced in the subsequent waves. Thus, we focus on the initial outbreak period, when the long-term consequences of the virus were still poorly understood, vaccines were still not available, and policy-makers were not certain which control measures would be effective. With this approach, we hope to suggest a method to infer the efficiency of restriction measures and to inform future urgent preparedness response plans in the time-critical phase of a pandemic.

We estimate the impact of individual NPIs on social distancing and Covid-19 spread using causal analysis methodology, taking into account confounding factors such as concurrent NPIs, Covid-19 morbidity measures, and country-level socio-economic and demographics factors. We tested two different variables as outcomes in the causal effect estimation. First, we analysed how NPIs influence the mobility trends across different categories of places such as residential and retail/recreation areas. In the second approach, we evaluated the effect on the growth rate of the reproduction number R_t_, i.e., the rate by which the pandemic spreads. To the best of our knowledge, this is the first study to quantify NPI effectiveness using a causal inference framework in such a wide geography coverage, while accounting for confounding biases and performing sensitivity analyses to assess the robustness of our findings.

## Materials and methods

### Data collection and pre-processing

#### Non-pharmaceutical interventions

We extracted the data on the restriction policies in 113 countries using the Worldwide Non-pharmaceutical Interventions Tracker for Covid-19 (WNTRAC) [20], a comprehensive database consisting of over 8, 000 Covid-19-related NPIs implemented worldwide. WNTRAC was updated periodically until October 5^th^ 2021, and we use the latest release for the analyses described hereby, which covers the period until the day before. For our experiments, we selected a subset of the five most well-defined and frequently imposed NPIs in the data: confinement, entertainment/cultural sector closure, work restrictions, mask-wearing, and school closure. Table 1 shows each NPI with its respective description.

**Table 1.** Description of the five NPIs used in this study.

| NPI | Description |
| --- | --- |
| School closure | Closing of any school category: kindergartens/daycares, primary/secondary schools or universities. |
| Cultural closure | Closing of any cultural establishment: bars, restaurants, night clubs, museums, theaters, cinema, libraries, festivities, parks and public gardens, gyms and pools, or churches. |
| Confinement | Physical distancing and universal lockdown measures. Considered as imposed when it is mandatory for the entire population. |
| Work-from-home order | Work restrictions and mandatory work-from-home orders for all nonessential workers. |
| Mask wearing | Facial coverings or mask wearing. Considered as imposed if it was mandatory or recommended in public spaces. |

### Mobility trends

To infer people’s dynamic behavioral response to restriction policies, we obtained Google mobility data [21]. These reports include the per-day change in movement across different categories of places compared to a baseline day before the pandemic outbreak. Similar to [17], we chose the change in duration of time spent in residential areas as a primary metric to measure social distancing and policy adherence. We additionally considered the effect of NPIs on the changes of movement in retail and recreation areas, generally seen as nonessential visits. For each category, we smoothed weekly patterns by using the seven-day rolling averaged mobility. When values were missing, we performed a linear interpolation. We used country-level information for all countries in our analysis, except for the US, which contained state-level data for both NPI and mobility data.

### Socio-economical and health indicators

We used development indicators compiled from officially recognized international sources to account for heterogeneity in terms of socio-economic and health factors within individual countries. From the World Bank’s World Development Indicators (WDI) [22], we selected a subset of variables, including access to electricity, outdoor air pollution, and forest area. We also included country-level health information, ranging from life expectancy at birth, smoking rate, and prevalence of undernourishment. As we have indicators per year, we take the most recent metrics available per country. Similar covariates were used in previous works [3, 4].

We also included variables describing population distribution, age-structure, and human development index from Our World in Data (OWID) [23]. To avoid producing highly biased estimates with missing values imputation, we only analysed countries with data available for at least 70% of the WDI and OWID variables and imputed the remaining missing features with the mean. We characterize each country with a cluster ID obtained through DBSCAN clustering [24] (scikit-learn, version 1.0.1) performed on the vector of socio-economical and health features (see Table 1S with the final set of features, and 1S Fig in Appendix E1 for a detailed explanation on the clustering method).

### Covid-19-related variables

Our model used daily cumulative confirmed cases and deaths from the WHO reports [25]. Before estimating R_t_, we smoothed the number of new cases over time with a local polynomial regression using a window size of 14 days and a polynomial degree of 1 to minimize the impact on the edges. The 14 day span (7 days forward and backward) was used to ensure that an equal number of weekdays was used for smoothing and to account for a time lag between exposure, testing, and documenting case reports. We used the R package EpiEstim developed by Cori and colleagues [26] and extended by Thompson et al. [27]. We chose a gamma distributed serial interval (mean, 3.96 days [SD, 4.75 days]; this interval was constant across periods and derived from previous epidemiological surveys on Covid-19 [28]), to estimate R_t_ and its 95% credible interval on each day via a 7-day moving average. EpiEstim was chosen due to its statistically robust analytical estimates and extensive use in the disease epidemiology literature.

Because estimating the serial interval distribution may not be possible in the early phase of an outbreak, or may be associated with significant uncertainty as countries still had to establish their documentation practices, we exclude the period before 100 cumulative cases of each country from our dataset. Therefore, the window of analysis for each country starts on the day the cumulative cases exceeded 100 and ends on June 1^st^ 2020. When concatenating all data sources, we found an intersection of 113 countries that were used in our final analysis. Countries like China, Iran and part of central African countries were excluded due to missing mobility data (see geography selection in Appendix E2).

### Statistical analysis

#### Outcome prediction

To investigate whether the observed variables (Covid-19 morbidity measures, socio-economical factors and status of NPIs) contained the predictive power for the outcomes of interest, we built prediction models to estimate R_t_ and the change in mobility. We split the dataset into two non-overlapping sets: 70% of samples used for training and 30% for validation. We used the gradient boosted trees algorithm with the XGBoost [29] package (version 1.2.0; https://xgboost.ai), with hyperparameters tuned using 5-fold cross validation on the train dataset, via a randomized grid search. The TreeExplainer from the SHapley Additive exPlanations (SHAP) [30] package was fit on the validation set to estimate the association and contribution of each feature to the XGBoost model.

We evaluated the accuracy of the predictions using the mean-squared error (MSE). To compare between the performance of different models at different scales, we normalized the estimates and the observed values in the test set before computing the MSE. The 95% confidence intervals (CI) were obtained with 100 bootstrapping iterations.

#### Potential outcome framework

We formulated causal effects in terms of the potential outcome framework [31]. Each country *i* at each point in time *t* is characterized by a feature vector *X*_*i,t*_, consisting of dynamic variables - the cumulative and new number of cases/deaths per million - and a static variable - a socioeconomic cluster ID, which is country-specific and constant over time. In addition, we included information regarding the status of the restrictions. This is a feature vector containing a set of binary features corresponding to whether other NPIs are in place at that time point. We denote 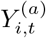 as the potential outcome in a given country *i* on day *t* for binary treatment *a*. We were interested in two possible outcomes: (i) 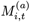, the difference in the country-level mobility defined as in [32] and (ii) 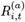, the change in SARS-CoV-2 transmission represented by a ratio of the reproduction number R_t_, defined as in [9]. We evaluated the outcomes with respect to a time-lag *w* (in days), as follows:

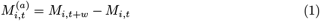

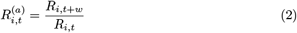

In this study, we considered the effect on the outcomes 2 weeks after the NPIs were enacted ± 1 week (i.e., w = 7, 14 and 21). Our goal was to estimate the Average Treatment Effect on the Treated (*ATT*), defined as

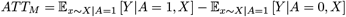

and

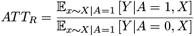

where *ATT*_*M*_ and *ATT*_*R*_ denote the causal effects of human mobility trends and R_t_, respectively. Thus, a null effect of NPIs on mobility would be equivalent to *ATT*_*M*_ = 0. Similarly, a null effect on R_t_ corresponds to *ATT*_*R*_ = 1.

To identify such a causal effect, we make several assumptions: (i) conditional exchangeability (ii) Stable Unit Treatment Value Assumption (SUTVA) and (iii) positivity. Identifying the ATT requires a weaker version of the assumptions above, as we are only estimating potential outcomes for the treated part of the population. A detailed discussion about the assumptions can be found in Appendix E3.

#### Study design

A crucial component for estimating causal effect from observational studies is the ability to conceptualize and to emulate randomized experiments. To this end, we designed a cohort study exclusively separating samples (in this case, days) from the treatment and control groups. For each NPI *k* ∈ *K*, we defined an *event* as the day NPI *k* was enacted in a certain country, given that this intervention was not in action the day before. We denote this day 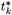. We defined the treatment group for NPI *k* as the set of 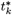 in the 113 countries during their respective period of analysis (Fig 1). The control group includes all days in the cohort except 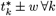 where *w* is the number of days before and after the event, and it is also the same time-lag period defined in Eq 1 and Eq 2. The control cohort represents a period where none of the five NPIs were enacted (an event did not occur), but they might still be in place during that period. Unlike the treatment groups, which are NPI-specific, the control groups are the same for all NPIs used as treatment.

**Fig 1.**
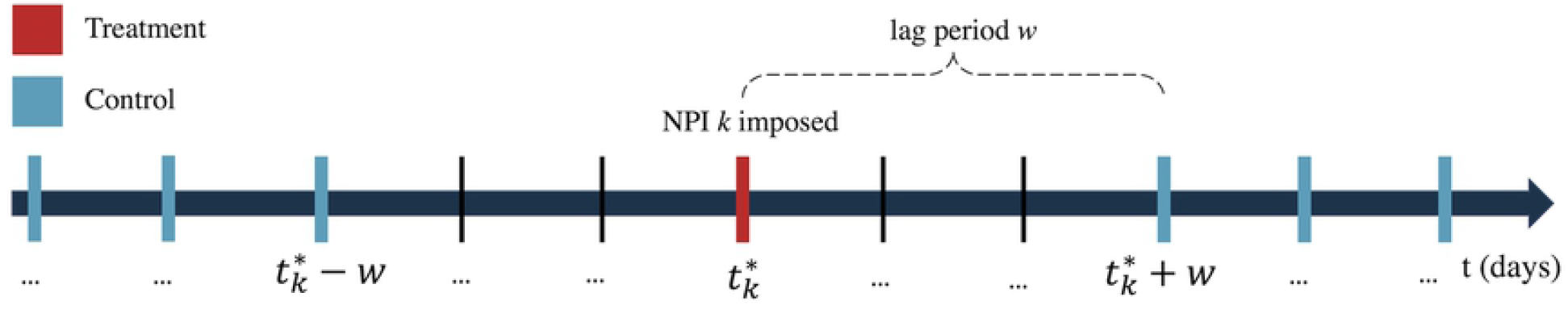
Schematic presentation of the study design. Vertical lines represent days. We denote day 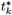 (in red) as the day when NPI *k* was enacted, considering that this NPI was not active the day before (i.e., event date). The treatment group for NPI *k* consists of 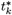 across all countries. The control group are the remaining days (in blue) outside the time interval 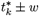, where *w* is a lag period.

#### Covariate balancing

We used Adversarial Balancing (AdvBal) [33] to estimate the mean potential outcome of the control group. In brief, AdvBal borrows principles from generative adversarial networks to assign weight to each sample in some source data, such that the resulting weighted data becomes similar to a given target data. Unlike the original paper, we defined only the treated group as the target population, and not the entire population, i.e., we fixed the weights of the treated samples and adjusted the weights of the control in order to balance the groups. The implementation of AdvBal and the resulting balancing evaluation of the models were done with the Python package *causallib* [34] (version 0.7.1; https://github.com/IBM/causallib). The final effects and the associated 95% confidence intervals were computed with 1, 000 bootstrap samples.

#### Sensitivity analysis

We performed sensitivity analyses to test our conclusions under different scenarios. First, we reran our framework with a different model to estimate treatment effect from observational data. We used inverse probability weighting (IPW) [35], a longstanding popular method that overcomes confounding by weighting samples by the inverse of their probability of being assigned to their treatment, conditioned on their covariates. These probability parameters were estimated with a logistic regression model. Once confounding was adjusted and the weights were computed, we estimated the causal effect on the treated sample as a weighted average of the outcome.

Second, we performed a complementary analysis to test whether the estimated effects were consistent when using an alternative study design. In this approach, the treatment groups are the same as in the original study (Fig 1). On the other hand, the control group consists of events from the remaining NPIs (apart from the NPI used as treatment). For example, if we were to estimate the effects of NPI *k* ∈ *K*, the treated group would contain only the events for this specific NPI, 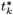, whereas the control group would be the events of all other NPIs K_-k_, as long as these events do not overlap with 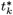.

## Results

### NPI employment statistics

As a preliminary step, we examined the frequency at which each NPI was imposed in the 113 countries (Fig 2). Since governments introduced and lifted lockdowns and social distancing measures consistently throughout time, we observed a continuous growth in the number of confinement events (Fig 2A). This behavior was the opposite for mask wearing events, which had a swift and sustained increase until October 2020, and then stayed constant for the rest of the period. By the end of March 2020, over 60% of all documented NPI events until October 2021 had already happened. We then analysed the distribution of NPIs across different countries before and after June 1^st^ 2020 (5S Fig in Appendix E7). We discovered that in the first wave, most countries imposed a large and diverse set of NPIs, which was greatly reduced in the following waves. For example, European countries like Spain and Italy introduced very similar government policies during their first waves, but adopted different strategies in the subsequent period: while Italy imposed similar preventive measures with less frequency, Spain placed more emphasis on lockdowns. Overall, throughout 2021, countries had to choose the appropriate NPIs that best fit their socio-economic circumstances and gradually lifted restrictions to avoid negative impacts in the economy [36].

**Fig 2.**
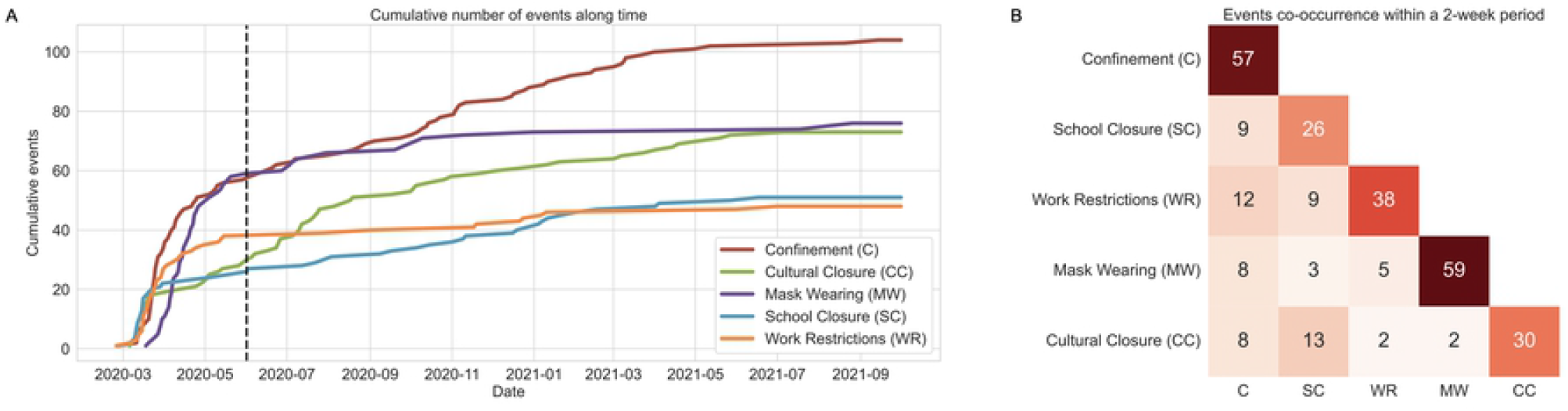
Distribution of NPIs and frequency of their co-existence. (A) Cumulative number of events over time. An event is defined as the day when an intervention was enacted in a country, considering that this NPI was not active the day before. Until June 1^st^ 2020 (dashed vertical line), 60% of all events had already happened. (B) Co-occurrence matrix of events in the limited period until June 1^st^ 2020. The diagonal values represent the number of events. Cell values indicate the sum of times that NPI *i* (x-axis) and NPI *j* (y-axis) were enacted together within a period of 2 weeks in the same country or US state.

Because a large number of restriction measures were initiated simultaneously in multiple countries within a short period of time, we analysed the events’ coincidence per country until end of May 2020. The co-occurrence matrix in Fig 2B shows that the NPIs are co-linear in this period, i.e., they frequently co-occurred. For example, out of 57 confinement events, 12 of them occurred within a period of two weeks (seven days earlier or later) of a work restriction event in the same country. When high co-linearity exists, individual effect estimates are more challenging, as it is harder to identify what in fact drives the change in SARS-CoV-2 transmission. To overcome this problem and to control for the influence of individual NPIs on the estimated effects, we opted to add the status of NPIs as confounders to our causal model (recall section Potential outcome framework).

### Predictors of mobility change and Covid-19 R_t_

The prediction performance on the validation dataset of both outcomes and their feature contribution plots are summarized in Fig 3. To evaluate the performance of the XGBoost models, we compared their accuracy to the one of a model based on permutation tests [37]. This model generates a null distribution by calculating the normalized MSE under the null hypothesis, where in each bootstrap replicate the features are kept the same but the outcomes undergo different permutations. We found that for all scenarios considered in this study (different time lags and different outcome variables), the features we utilized showed enough statistical power to allow a good prediction (Fig 3C). Associations between the outcome variables and baseline features can be visualized using SHAP “beeswarm” plots, that show the top-10 contributing features for the outcomes prediction using a time-lag of 7 days (Fig 3A-B). The strongest predictor of R_t_ was its own value 7 days before, implying a clear positive association between R_t_ measures within a one-week period (Fig 3A). Such strong a association was not observed with other important features, such as confirmed cases and deaths. Changes in mobility within several categories also impacted the R_t_ prediction. In particular, an increase in mobility in workplaces and retail contributed to a higher SARS-CoV-2 transmission. The feature importance analysis of residential mobility prediction revealed that mobility categories are highly correlated: as the time spent at home increases, the movement outside residence decreases (Fig 3B).

**Fig 3.**
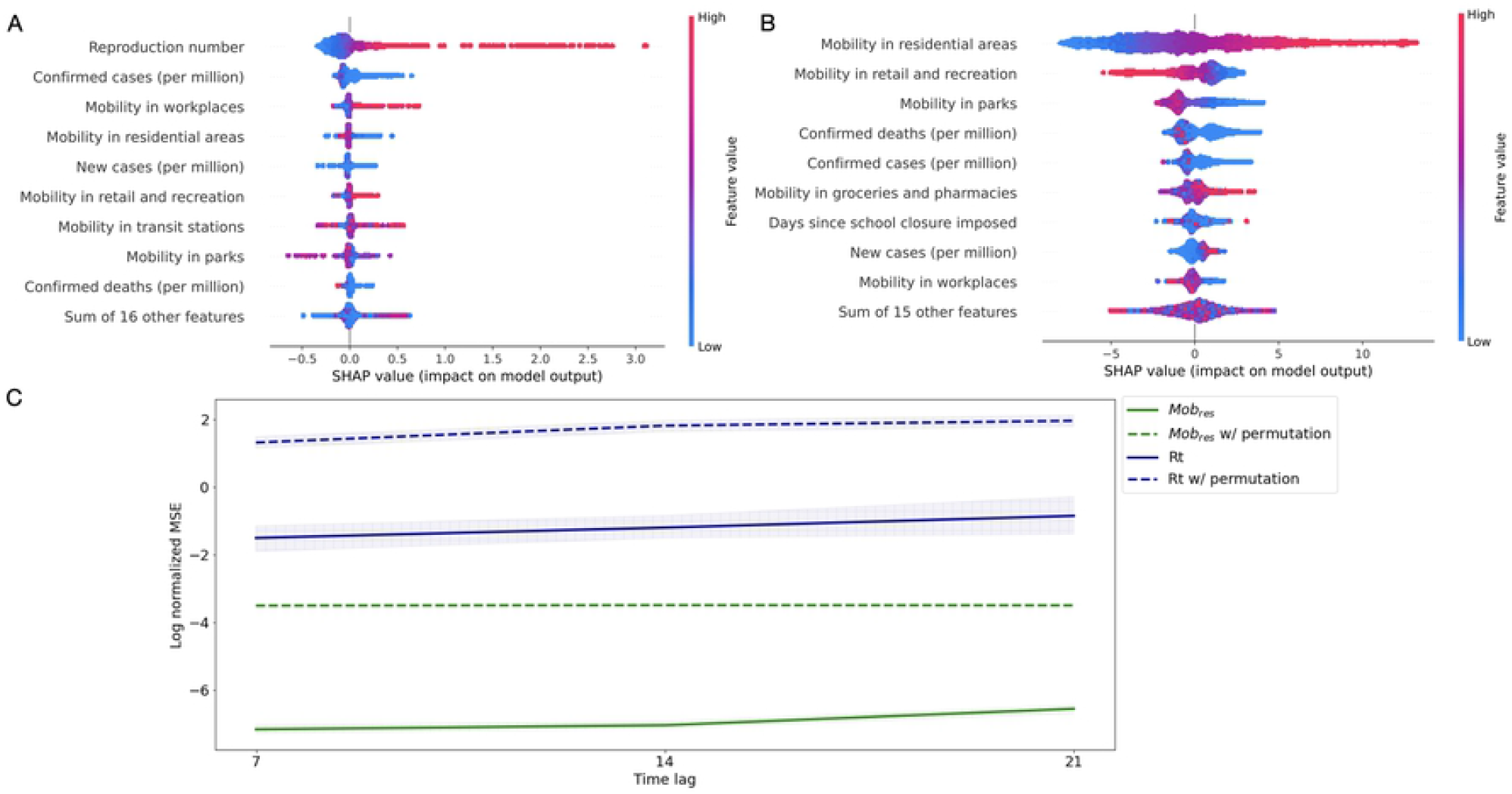
Important predictors of outcome and accuracy of prediction for different time-lags. (A) Identification of top-10 predictive variables affecting R_t_ estimation. (B) Top-10 most contributing features for residential mobility estimation. SHAP analysis in A and B is based on predictions using a time-lag of 7 days. Each dot represents a single data sample in the validation set (i.e., country at a date). The dot color represents the feature value (red=high, blue=low). The farther a dot is from 0 on the x-axis, the more effect (positive or negative) this feature had on the prediction model for this particular sample. (C) Model performance (log of normalized MSE) of R_t_ (solid dark blue line) and residential mobility (solid green lines) in the validation set for different time lags. The dashed lines of the same color correspond to random prediction derived by permutation tests with their respective models. Both models significantly outperformed random prediction.

### Estimated effect of interventions on policy adherence and SARS-CoV-2 transmission

We derived causal effect estimates using balancing weights that minimized the confounding biases. The absolute standard mean difference (ASMD) was used as a measure to compare the distribution of observed baseline covariates between treated and untreated groups. The AdvBal algorithm could significantly decrease differences between covariate distributions, bringing the ASMD to less than 0.25 for all covariates (2S Fig in Appendix E4). This value is considered a reasonable cutoff for acceptable standardized biases, indicating that the effect estimates are reliable and robust to confounding [38].

We report the estimated causal effects 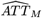 and 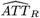 without balancing weights (unadjusted) and with balancing weights generated by the AdvBal algorithm and IPW (Fig 4). All estimates are presented with 95% confidence intervals and were obtained after assessing whether the balancing weights approaches were able to reduce biases.

**Fig 4.**
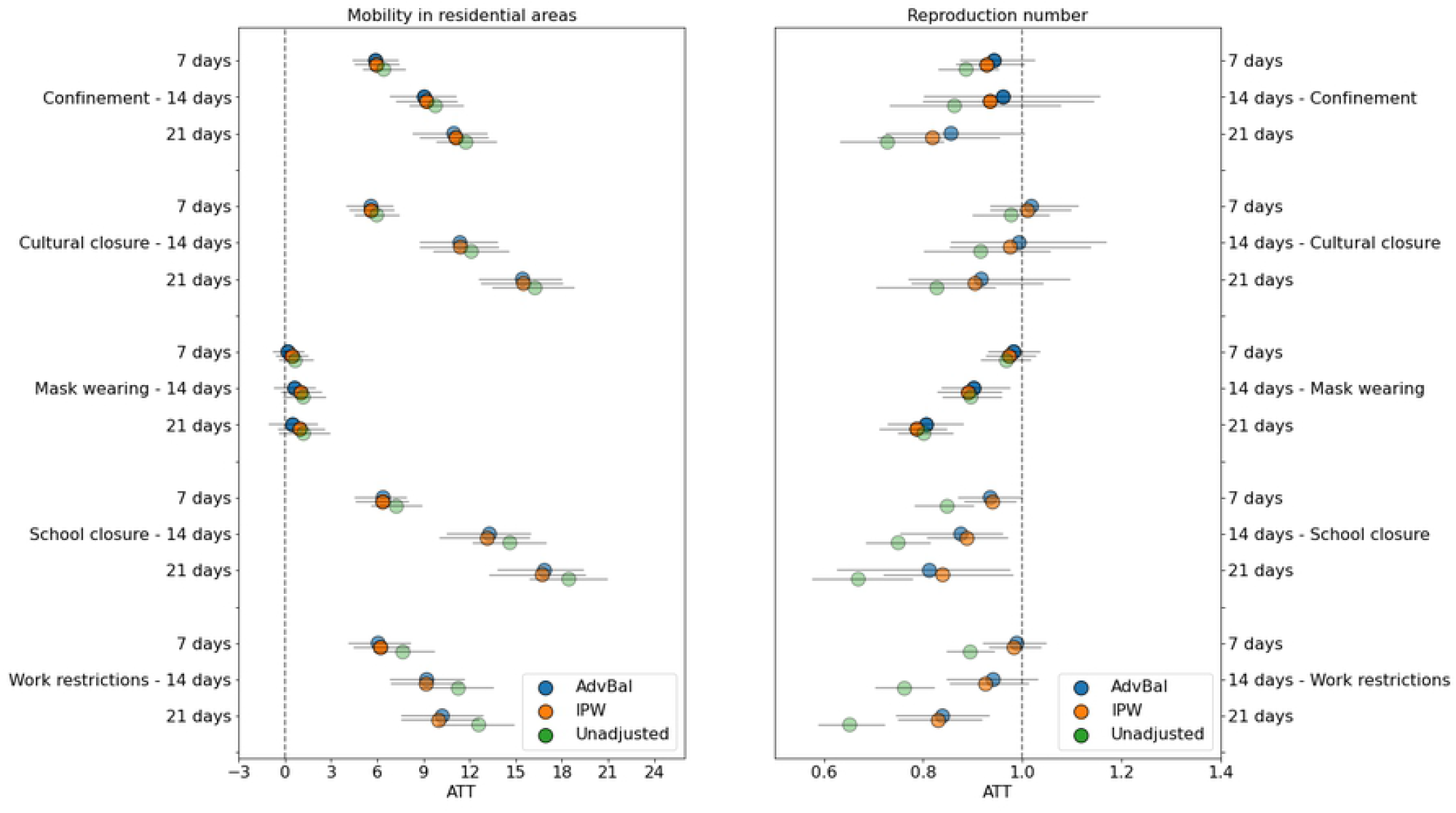
Estimated causal effects of NPIs over time on residential mobility (left) and reproduction number rate (right). In both plots, the opacity of the markers represents the ability of the balancing weights method to balance the treatment groups: the smaller the ASMD is, the more opaque the markers are. Fully opaque markers indicate an ASMD *<* 0.1, half-transparent markers indicate 0.1 ≤ ASMD ≤ 0.25 and most transparent ones represent ASMD *>* 0.25. Apart from mask wearing mandates, all NPIs caused a significant increase in time spent at home. Of those, school and cultural closures were the most effective. Closing schools, issuing face mask usage, and work-from-home mandates also caused a persistent reduction in R_t_ after their initiation, which was not observed with the other social distancing measures. Code used for generating figure is available at https://github.com/barakm-ki/symptoms-dynamics-of-COVID-19-infection/blob/master.

Our results suggest that the full extent of NPIs, except mask wearing, significantly affected human mobility change as early as 7 days after their initiation. However, changes in mobility varied between these 4 restrictions. School and cultural closures caused a quick and sustained increase in the time spent inside the home, whereas confinement and work-from-home orders had a slower and plateauing effect over time. Estimates from the AdvBal algorithm suggest that, following the introduction of NPIs, the time spent at home 14 days later was estimated to increase by 9.06% [95% CI: 6.86%, 11.10%], 9.22% [95% CI: 6.90%, 11.66%], 13.29% [95%CI: 10.57%, 15.95%] and 11.38% [95%CI: 8.83%, 13.80%], in response to confinement, work restrictions, school and cultural closure, respectively. Since people already spent a lot of time in their residence before the pandemic, movement changes in residential areas were likely to be smaller compared to outside locations, such as recreation and retail areas (3S Fig in Appendix E5).

From the five interventions considered, all except cultural closure and confinement caused a significant decline in the transmission of SARS-CoV-2, but within different time lags. It took an estimated 7 days after initiation of school closures to bring *A*-*T T*_*R*_ below 1, as opposed to 14 days for face mask mandates, and 21 days for work restrictions. By the end of 3 weeks, school closures had continuously reduced R_t_ until 0.81 [95% CI: 0.63, 0.98], mask wearing caused a decrease until 0.81 [95% CI: 0.73, 0.88], and work-from-home orders led to R_t_ reduction of 0.84 [95% CI: 0.75, 0.93]. Overall, the confidence intervals for the individual effects of all restrictions overlapped, suggesting a comparable effect between each other. IPW and the AdvBal algorithm showed similar trends in their resulting effects, but the balancing of covariates was marginally better on the latter (2S Fig, Appendix E4).

Results of a complementary analysis based on an alternative study design are described in Appendix E6. We found that under a stricter cohort design, where we compare the effect of NPIs with respect to the other NPIs, school closure had the greatest impact on increasing mobility in residences and in reducing R_t_ 14 days after its initiation (4S Fig).

## Discussion

We constructed a dataset that combines rich information about countries and their reaction to the urgent need to control the pandemic spread. The data include information on social-economics and health benefits, NPIs, and mobility data from more than 100 countries. We showed that the data have predictive power, and that the prediction of changes in mobility after imposing NPIs is more accurate than the prediction of the reproduction number. We employed a causal inference approach to quantify the effect of NPIs on the rate of R_t_ (i.e., transmission of SARS-CoV-2) and on the change of human mobility, which is considered a proxy measure of population adherence and social distancing. The purpose of this study was to help infer the efficiency of interventions in the early months of a pandemic, when a number of control measures had already been imposed by multiple countries in the absence of vaccines.

The most common use of causal inference seeks to estimate the average treatment effect (ATE). Such an analysis would answer questions such as “what would have happened if every country applied the NPI?”. However, in this case an analysis of this type proved to be unfeasible, since it was not possible to balance the confounding differences between the treated and untreated countries. We therefore opted to measuring the impact of NPIs in the countries that chose to impose the NPIs, which is known as the average effect on treated (ATT). This answers the question of “what was the effect of applying the NPIs in the countries that applied it?”. This approach allowed covariate balancing, which provided more reliable estimates of the effect of NPIs.

Our findings showed that mask wearing did not significantly impact mobility patterns in the first wave. Although a number of countries favored face mask usage early in their outbreaks [39], the main reason people changed their behavior was social distancing policies. We found that issuing confinement, work-from-home orders, or school/cultural closure mandates resulted in high levels of policy compliance even one week after their initiation, as measured by changes in movement in residential areas. This result was consistent with recent findings [32, 40].

The estimated effect on R_t_ showed that not all NPIs significantly contributed to a decrease in SARS-CoV-2 transmission. In particular, school closure achieved a sustained decline on the rate of R_t_, similar to what was found in observational studies of the first wave [4, 5, 7–9] and second European wave [41]. Because infected children can experience mild or no symptoms more frequently than older individuals [42] and tend to have more social contact than adults [43], it is expected that closing schools would considerably contribute to reduce the transmission. In contrast, on its own, closing cultural establishments does not seem to have an effect on the reproduction number, nor do work restrictions in the first 14 days after imposing the NPIs. Notably, we did not find substantial differences in the results when performing sensitivity analysis.

Our study extends previous first-wave estimate studies [2, 4, 5, 7–10] in a number of ways. First, we used the potential outcome framework to infer NPI effects. In its simplest form, our causal model made use of standard causal inference methods to correct observed biases and obtain valid effects with more transparent confidence intervals. Second, we addressed the issue of concurring NPIs by using their status as covariates in the causal model. By ensuring that all NPI-related covariates are well balanced between treatment groups, we enhanced the power to detect independent NPI effects. Third, we account for heterogeneity of countries by including a social economical cluster indicator in the dataset. Fourth, we conducted complementary analyses under alternative scenarios to test our conclusions not only with a different cohort study design, but also with another balancing weights generation approach.

We acknowledge several limitations in our analyses. Even with data from multiple countries that had diverse sets of interventions in place, inferring NPI effects still remained a challenging task. First, the R_t_ estimation was based on epidemiological parameters that are only known with uncertainty, due to many mild or asymptomatic cases that make it difficult to model the timing for the onset of symptoms and serial interval distributions. On top of that, R_t_ also relies on the data of confirmed cases, which were generally unreliable in the early days of the pandemic due to lack of testing availability and not-established documentation practices. To account for this, we began our analysis at each country’s 100^th^ case. Secondly, the data are retrospective and observational, meaning that unobserved factors could confound the results. Third, we were unable to assess the effect of lifting interventions. Since the NPI events in WNTRAC are automatically extracted from Wikipedia articles, which report the introduction of NPIs more frequently than their relaxation, the number of lifting events documented in the database did not have enough statistical power for causal inference. Yet, we believe we set the ground for a thorough analysis of NPIs and we were able to draw conclusions regarding the effect different NPIs had on the pandemic spread. Future work can assess the causal effect of the post-vaccine newly defined NPIs where health certificate notions were introduced in somewhat similar ways across different countries.

## Data Availability

The NPI data underlying the results presented in the study are publicly available in the WNTRAC repository at https://github.com/IBM/wntrac/tree/master/data. Google mobility data is available at https://www.google.com/covid19/mobility. Socio-economical and health indicators were collected from the World Bank database at https://databank.worldbank.org/source/world-development-indicators and from Our World in Data (OWID) at https://ourworldindata.org. Finally, the data on Covid-19 morbidity measures, such as confirmed cases and deaths, were downloaded from the World Health Organization (WHO) reports available at https://covid19.who.int/info

https://www.google.com/covid19/mobility

https://databank.worldbank.org/source/world-development-indicators

https://ourworldindata.org

https://covid19.who.int/info

## Code availability

The source code for the causal inference evaluation of NPIs is available in a public GitHub repository at https://github.com/IBM/causallib. Please refer to the README file in the repository for further instructions on using the code. Requests for the code used to generate the results and the plots should be directed to the corresponding author.

## Supplemental material

### Appendix E1

#### Clustering of countries based on socio-economic and health variables

We characterized each country by a cluster ID representing its socio-economic and health status. We used DBSCAN (Density-Based Spatial Clustering of Applications with Noise) [24] because the number of clusters generated was not limited to a pre-defined constant. The parameters *eps* and *Minpts* were decided based on experimental trials. First, we arbitrarily set five countries to be the minimum number of samples in a neighborhood and used the Euclidian metric to calculate distances between instances. The parameter *eps*, i.e., the maximum distance two countries can be from one another while still belonging to the same cluster, was selected according to a k-dist graph [44]; this graph calculates the distance between a point and its k-th nearest points (in this case, k = 5). We then plotted the distances in ascending order on a k-distance graph and chose the value at the point of maximum curvature, where the graph has the highest slope. Finally, noise samples were manually assigned to the defined clusters using domain knowledge. This procedure resulted in four clusters (1S Fig). Finally, we one-hot encoded the cluster identifier and represented the socioeconomic status with four binary variables: cluster 1, cluster 2, cluster 3, and cluster 4. The list of features used in the DBSCAN algorithm are detailed in 1S Table with their descriptive statistics.

**1S Fig. Clusters performed on socio-economic and health variables**. Countries are colored by their respective cluster ID. The DBSCAN algorithm found a total of 4 clusters: a large group comprising mostly African, South/Central American, and South East Asian countries (in dark green); another cluster predominantly with Eastern European countries (light green); the third cluster with the USA, Canada, and part of Western Europe (light orange); and lastly, a group of countries composed of the UK, along with a few Western European and Asian countries (dark orange).

### Appendix E2

#### Geography selection

The geography was selected based on the intersection of countries with available NPI data, mobility trends and at least 70% non-missing variables from socioeconomical and health databases (World Development Indicators [WDI] and Our World in Data [OWID]). The final dataset consisted of the following 113 countries:

Afghanistan, Angola, United Arab Emirates, Argentina, Antigua and Barbuda, Australia, Austria, Belgium, Burkina Faso, Bangladesh, Bulgaria, Bahrain, Bahamas, Bosnia and Herzegovina, Belarus, Belize, Bolivia, Brazil, Barbados, Canada, Switzerland, Chile, Colombia, Cabo Verde, Costa Rica, Czechia, Germany, Denmark, Dominican Republic, Egypt, Spain, Estonia, Finland, Fiji, France, Gabon, United Kingdom, Georgia, Ghana, Greece, Guatemala, Honduras, Croatia, Hungary, Indonesia, India, Ireland, Iraq, Israel, Italy, Jamaica, Jordan, Japan, Kenya, Kyrgyzstan, Cambodia, Republic of Korea, Kuwait, Laos Peoples Democratic Republic, Lebanon, Libya, Sri Lanka, Lithuania, Luxembourg, Latvia, Morocco, Moldova, Republic of, Mexico, North Macedonia, Mali, Malta, Myanmar, Mongolia, Mauritius, Malaysia, Nigeria, Netherlands, Norway, Nepal, New Zealand, Oman, Pakistan, Panama, Peru, Philippines, Papua New Guinea, Poland, Portugal, Paraguay, Qatar, Romania, Russian Federation, Rwanda, Saudi Arabia, Senegal, Singapore, Serbia, Slovakia, Slovenia, Sweden, Togo, Thailand, Trinidad and Tobago, Turkey, Ukraine, Uruguay, United States, Venezuela, Viet Nam, Yemen, South Africa, Zambia, Zimbabwe.

**1S Table.** Descriptive statistics for observed variables from OWID and the World Bank’s WDI datasets (accessed on October 10, 2021) across 113 countries.

| Variable | Mean ± std | Observations |
| --- | --- | --- |
| **Socio-economic variables** |  |  |
| Access to electricity | 92.36 ± 17.03 | % of population |
| Access to electricity | 97.43 ± 6.25 | % of urban population |
| Forest area (%) | 30.92 ± 21.25 | land in hectares |
| Air pollution, mean annual exposure | 26.91 ± 20.50 | micrograms per cubic meter |
| GDP per capita | 23198.36 ± 20778.14 | |
| Human Development Index | 0.76 ± 0.13 | |
| People with basic handwashing facilities | 59.93 ± 17.64 | % of population |
| Hospital beds | 3.32 ± 2.49 | per 100,000 people |
| Population density | 234.21 ± 768.68 | |
| Median age | 33.05 ± 8.70 | |
| Population ages 65 and above | 10.33 ± 6.65 | % of total population |
| Population ages 70 and above | 6.60 ± 4.57 | % of total population |
| **Health variables** |  |  |
| Cardiovascular death rate | 242.38 ± 120.20 | per 100,000 people |
| Life expectancy at birth, total | 74.59 ± 6.45 | in years |
| Antiretroviral therapy coverage | 60.39 ± 14.23 | % of people living with HIV |
| Smoking prevalence, males | 32.21 ± 17.64 | % of adults |
| Smoking prevalence, females | 12.18 ± 11.39 | % of adults |
| Prevalence of undernourishment | 7.33 ± 9.88 | % of population |
| Prevalence of overweight | 50.74 ± 6.73 | % of adults |
| Cause of death by communicable diseases and maternal, prenatal and nutrition conditions | 16.45 ± 9.58 | % of total |
| Cause of death by non-communicable diseases | 74.91 ± 18.31 | % of total |
| Diabetes prevalence | 7.8 ± 3.97 | % of population ages 20 to 79 |
| Mortality from CVD, cancer, diabetes or CRD | 17.72 ± 5.82 | % of ages 30 to 70 |

Because we had more locally fine-grained mobility and NPI data available for the United States, we used the state-level information of all 50 states.

### Appendix E3

#### Identification assumptions

The Rubin Causal Model [31] used in this study is an approach to causal inference that is based on a framework of potential outcomes. Under this framework, causal effects are estimated with comparisons of potential outcomes under the two different treatments: one that received the intervention, and another that received a different intervention (e.g., placebo or no treatment). In our context, these two groups are defined as days in countries where NPIs were imposed vs. days in countries where NPIs were not imposed. Causal estimates from this source are unbiased when three assumptions are met, namely (i) conditional exchangeability, (ii) Stable Unit Treatment Value Assumption (SUTVA) and (iii) positivity.

Conditional exchangeability states that the counterfactual outcomes are conditionally independent of the treatment given the set of covariates. Under this assumption, the treatment groups are “exchangeable”, i.e., there are no unmeasured confounders that are a common cause of both treatment and the outcome [45]. In other words, if all confounders are measured, then we can assume that exchangeability holds within the strata dictated by the confounders, and we can estimate the causal effect by using methods that eliminate the confounding (e.g.,, by emulating a randomized controlled trial [RCT] using balancing weights methods). This is because randomization ensures that the covariates associated with the outcome are equally distributed between the treatment groups. In reality, in observational studies as the one we studied here, one cannot empirically verify that conditional exchangeability holds or there is no unmeasured confounding. Thus, causal inference relies on subject-matter knowledge to identify possible confounders in the data, so that the assumption is at least approximately true.

SUTVA stipulates that the outcome of one unit should not be affected by another unit’s treatment assignment. Although SUTVA plays a central role in the identification of causal effects, this assumption does not hold in many settings. For example, when a certain country bans inbound flights from its neighbouring countries (introduction of a new NPI), the observable outcome (mobility change, R_t_ rate or any other Covid-19 morbidity measure) of the banned countries are certainly affected by this treatment assignment. Another classical example is given in epidemiology, where the possibility of an individual becoming infected depends on whether the population is vaccinated. In our study of NPI effectiveness, SUTVA is an unrealistic assumption. Yet, we claim that our final estimated effects are still a good enough approximation. We direct the reader to the work of Hudgens and Halloran [46]. In their paper they do a review of previous studies that estimated causal effects in the presence of interventions’ interference.

Positivity, also called a lack of covariate overlap, is the assumption that any sample has a positive probability of receiving all values of the treatment variable. In other words, to identify causal effects pertaining to a treatment A, there must be some probability of receiving A given a certain baseline of covariates X, otherwise treatment versus control causal effects cannot be identified. Mathematically, *Pr*(*A* = *a*|*X* = *x*) *>* 0 for all *x* where *Pr*(*X* = *x*) ≠ 0. The positivity assumption has the very important consequence of ensuring that features in both treatment groups are equal in their distribution. In our work, we empirically verify to some extent whether positivity holds by checking whether the distribution of covariates is similar between the two treatment groups. By inspecting the success of the weighting method used (in our case, either IPW or the AdvBal algorithm [33], we are ensuring that positivity holds in our dataset. In the next section, we discuss the balancing evaluation of the applied weights in more detail.

### Appendix E4

#### Balancing evaluation

A fundamental step required to produce reliable effect estimates is to control for systematic differences between the treatment and control groups. To this end, balancing weights methods, such as adversarial balancing (AdvBal) [33], generate weights that reduce the observed confounding biases, and thus can be used to emulate a randomized controlled trial (RCT) by re-weighting the population. To evaluate the performance of AdvBal, we analysed the distribution of all covariates across the treatment groups before and after re-weighting samples with AdvBal. The covariate balancing plot in 2S Fig is an example of an evaluation plot when work restrictions was assigned as treatment. In the plot, the difference in distribution between treatment groups is quantified by the absolute standard mean difference (ASMD), defined as the absolute value in the difference in means of a covariate between the treatment groups, divided by its standard deviation in the treated group. A small ASMD value represents a good balance, while a value larger than some threshold is considered imbalanced. In this study, we considered a threshold of 0.1 as a reasonable cutoff for acceptable ASMD [38]. We observed that before balancing, many features were biased between the treated and untreated groups. AdvBal was able to minimize this discrepancy, bringing the ASMD to less than 0.1 for all covariates.

**2S Fig. Balancing evaluation plots of AdvBal algorithm (upper) and IPW (lower) of the dataset during train phase**. The plot displays the absolute standard mean difference (ASMD) of each feature in the original unweighted data (orange triangles) and in the weighted data (blue circles) obtained with the weights generation method. Although both methods were able to balance all covariate distributions and bring ASMD below 0.1, AdvBal performed marginally better.

### Appendix E5

#### NPIs effect on places of recreation

The mobility data for retail and recreation areas represents the change (relative to the period before the pandemic) in the number of visitors to places like restaurants, shopping centers, and libraries. The estimated causal effects show that, apart from mask wearing, all social distancing policies were effective in decreasing visits to this category of places (3S Fig). Of these, school and cultural mandates were the most effective ones, achieving a 50.4% [95% CI: 41.9%, 58.2%] and 47.4% [95% CI: 39.4%, 55.1%] average reduction in 21 days, respectively. Overall, changes in mobility outside residential areas were larger in magnitude.

**3S Fig. Effect of NPIs over time on retail and recreation areas**. Model results for the 113 countries show that school and cultural mandates were the most effective ones, achieving a 50.4% and 47.4% reduction in the average number of visitors to recreation areas in 21 days, respectively. AdvBal: Adversarial balancing algorithm, IPW: inverse propensity weighting.

### Appendix E6

#### Complementary analysis of effect of NPIs

In an alternative cohort study design, the treatment group is composed of all events of a certain NPI of interest, whereas the control group contains all events of the remaining NPIs. Thus, instead of estimating the effect of NPIs compared to a period when no NPIs are imposed, we aimed at estimating the effects of individual NPIs relative to others. We consider this scenario a more “strict” design where we hoped to examine whether our findings would be consistent with the results of the original study.

Results of this complementary analysis are shown in 4S Fig. We found that compared to other NPIs, school closure was the most effective restriction in changing the two mobility categories 14 days later after its initiation. It was also the only NPI that greatly impacted R_t_ in the same time period.

**4S Fig. Estimated causal effects of NPIs under the study design “NPI vs. other NPIs”**. Under this approach, we investigated which NPI has the highest impact compared to the others. AdvBal: Adversarial balancing algorithm, IPW: inverse propensity weighting.

### Appendix E7

**5S Fig. NPI employment statistics per country and US state before and after June 1**^**st**^ **2020**. Until June 1^1st^, governments worldwide enacted restrictions in a more consistent way, i.e., the distribution of the NPIs employed was more similar across countries.

